# *LRRK2* in Focus: A Global Browser Linking Genetic Diversity to Functional Effects

**DOI:** 10.64898/2026.07.01.26357034

**Authors:** Spencer M. Grant, Vesna van Midden, Elias Fernandez-Toledo, Momodou Cham, Esther Sammler, Dario R. Alessi, the Global Parkinson’s Genetics Program, Huw R. Morris, Cornelis Blauwendraat, Andrew B. Singleton, Lara M. Lange

**Affiliations:** Center for Alzheimer’s and Related Dementias (CARD), National Institute on Aging and National Institute of Neurological Disorders and Stroke, National Institutes of Health, Bethesda, MD, USA; Department of Clinical Neurosciences, School of Clinical Medicine, The University of Cambridge, Cambridge, UK; University Medical Centre Ljubljana, Ljubljana, Slovenia; University of Concepción, Concepción, Chile; Richard Novati Catholic Hospital, Sogakope, Ghana; Division of Neuroscience, Faculty of Health, University of Dundee, Dundee, UK; Medical Research Council Protein Phosphorylation and Ubiquitylation Unit, Faculty of Life Sciences, University of Dundee, Dundee, UK; Department of Clinical and Movement Neurosciences, UCL Queen Square Institute of Neurology, London, UK; UCL Movement Disorders Centre, University College London, London, UK; Global Parkinson’s Genetics Program (GP2), Chevy Chase, MD, USA; Coalition for Aligning Science (CAS), Chevy Chase, MD, USA; Laboratory of Neurogenetics, National Institute on Aging, Bethesda, MD, USA; Institute of Neurogenetics, University of Luebeck, Luebeck, Germany

## Abstract

**Background:** *LRRK2* variants are major contributors to Parkinson’s disease (PD). Many pathogenic variants increase kinase activity, underscoring the value of functional assays in nominating therapeutic targets and kinase inhibitors as potential disease-modifying therapies.

**Objective:** To develop an interactive resource that provides functional context and ancestry-specific variant frequencies.

**Methods:** Genotyping and short-read sequencing data were analyzed for 101,678 individuals (61,709 PD, 39,969 controls) from the Global Parkinson’s Genetics Program (GP2) and integrated with clinical and in-vitro biochemical kinase activity information.

**Results:** The *LRRK2* Browser (http://gp2.org/lrrk2browser) displays ancestry-specific genetic data for 19,596 *LRRK2* variants (968 exonic, 14 disease-associated) across 11 populations, and functional data for 171 variants. Clinical annotations include age, age at onset, and family history of PD.

**Discussion:** The publicly available *LRRK2* Browser represents an open-access, multi-ancestry resource to support *LRRK2* variant interpretation. It aims to enhance the translational potential of genetic and functional data for precision medicine and the implementation of gene-targeted therapies in diverse populations.

## Introduction

Variants in the *Leucine-rich repeat kinase 2 (LRRK2)* gene are major contributors to both monogenic and complex forms of Parkinson’s disease (PD), including rare, pathogenic variants with mostly incomplete penetrance as well as more common risk alleles. Initially identified in familial PD cohorts (1,2) and later supported by genome-wide association studies (GWAS) (3,4), altogether, more than 1,000 missense variants have been reported in *LRRK2* (5–8). However, only a small proportion have been confidently classified as disease-causing (9).

*LRRK2* encodes a large multidomain protein with GTPase and kinase functions that regulate key cellular processes, including endolysosomal vesicular trafficking, autophagy, cytoskeletal dynamics, immune signaling, and cellular stress pathways, which are increasingly implicated in neurodegeneration (10). Pathogenic *LRRK2* variants are thought to act primarily through a toxic gain-of-function mechanism, most notably through increased kinase activity (10). This knowledge has positioned LRRK2 as a leading target for disease-modifying therapies, with multiple kinase inhibitors and LRRK2-lowering strategies currently in different stages of clinical development (11,12) for PD treatment. Functional assays, particularly those measuring phosphorylation of substrates such as RAB10, together with segregation and population data, have enabled the classification of newly identified *LRRK2* variants as pathogenic, which is highly relevant for identifying individuals who may be eligible for targeted clinical trials (13–18).

A major consideration, especially related to *LRRK2* variant interpretation, is the marked variability in the frequency and spectrum of *LRRK2* variants across populations. For instance, p.G2019S is enriched in Ashkenazi Jewish and North African Berber populations, and p.L1795F has been identified exclusively in European populations (14). In contrast, p.G2385R, p.R1628P, and p.R1067Q are particularly relevant in East Asian populations (16,19).

Here, we present the *LRRK2* Browser (http://gp2.org/lrrk2browser) as a resource designed to address the current uncertainty surrounding the functional effects of many *LRRK2* variants, particularly within ancestry-specific contexts, which is essential for accurate interpretation. This interactive platform aggregates, visualizes, and contextualizes *LRRK2* variant data, integrating functional evidence, clinical annotations, and ancestry-specific allele frequencies. The browser is built using ancestrally diverse datasets from the Global Parkinson’s Genetics Program (GP2; https://gp2.org), a large international collaborative dedicated to defining the genetic architecture of PD through inclusive, large-scale population studies (20–23). This resource will serve as a valuable tool to support research, enhance diagnostic precision, and promote more equitable inclusion in LRRK2-targeted clinical trials and emerging therapeutic strategies.

## Methods

### Data extraction

Individual-level genetic and clinical datasets were obtained from GP2 Release 11 (https://doi.org/10.5281/zenodo.17753486). Genetic data processing, quality control (QC), and genetic ancestry predictions were performed according to GP2 standards using Genotools, as previously described (24). *LRRK2* variants were extracted from all available genetic datasets, including raw and imputed genotyping data generated with the Illumina NeuroBooster Array (NBA), clinical exome sequencing (CES), and whole-genome sequencing (WGS). Related individuals were removed for downstream analyses. The dataset included a total of 101,678 individuals; 61,709 were individuals with PD, and 39,969 were healthy, unrelated controls. The numbers of included participants stratified by phenotype, genetic dataset modality, and ancestry, are summarized in **Table 1**.

**Table 1:** Number of case and control samples for each data modality across each genetic ancestry.

| **Ancestry** | **Phenotype** | **WGS** | **NBA** | **CES** | **Total\*\*** |
| --- | --- | --- | --- | --- | --- |
| **AAC** | PD Cases | 230 | 475 | 147 | 535 |
|  | Healthy Controls | 158 | 836 | 0 | 871 |
| **AFR** | PD Cases | 1315 | 2556 | 96 | 2624 |
|  | Healthy Controls | 2060 | 4224 | 0 | 4307 |
| **AJ** | PD Cases | 1149 | 2135 | 800 | 2646 |
|  | Healthy Controls | 228 | 761 | 0 | 870 |
| **AMR** | PD Cases | 794 | 2100 | 320 | 2675 |
|  | Healthy Controls | 301 | 1467 | 0 | 1611 |
| **CAS** | PD Cases | 209 | 769 | 38 | 852 |
|  | Healthy Controls | 117 | 399 | 0 | 418 |
| **CAH** | PD Cases | 509 | 979 | 398 | 1037 |
|  | Healthy Controls | 953 | 1313 | 0 | 1414 |
| **EAS** | PD Cases | 1431 | 3209 | 94 | 3356 |
|  | Healthy Controls | 1020 | 2678 | 0 | 2696 |
| **EUR** | PD Cases | 10682 | 32721 | 5802 | 38354 |
|  | Healthy Controls | 2522 | 24930 | 0 | 26016 |
| **FIN** | PD Cases | 28 | 106 | 16 | 119 |
|  | Healthy Controls | 6 | 19 | 0 | 21 |
| **MDE** | PD Cases | 786 | 933 | 42 | 1195 |
|  | Healthy Controls | 965 | 1286 | 0 | 1317 |
| **SAS** | PD Cases | 108 | 366 | 79 | 419 |
|  | Healthy Controls | 333 | 392 | 0 | 428 |
| **Other\*** | PD Cases | 0 | 0 | 6723 | 6723 |
|  | Healthy Controls | 0 | 0 | 0 | 0 |
| **Total** | PD Cases | 17241 | 46349 | 14555 | 61709 |
|  | Healthy Controls | 8663 | 38305 | 0 | 39969 |

WGS Whole-genome sequencing NBA NeuroBooster Array genotyping CES Clinical-exome sequencing AAC African-admixed ancestry AFR African ancestry AJ Ashkenazi Jewish ancestry AMR Admixed American / Latino ancestry CAS Central Asian ancestry CAH Complex Admixture History EAS East Asian ancestry EUR Non-Finnish European ancestry FIN Finnish ancestry MDE Middle Eastern ancestry SAS South Asian ancestry
\* Samples that only underwent clinical exome sequencing do not have genetic ancestry predictions.
\*\* Data are available from multiple modalities for some individuals.

Variants of interest (*LRRK2*, chr12:40224997-40369285; https://www.ncbi.nlm.nih.gov/gene/120892) were extracted using PLINK v1.9 and v2.0. In addition to GP2 QC metrics, imputed variants were filtered to only include those with an imputation quality score ≥0.8. Variants were annotated using ANNOVAR (25) using refGene, ClinVar (2025-07-29 release), dbNSFP v4.7a, gnomAD v4.1 (WGS), and dbSNP151 with allelic splitting. On the variant carrier level, additional data including family history status, age at onset (AAO) for individuals with PD, and age for unaffected variant carriers, were extracted where available.

### Functional LRRK2 kinase activity assay

We leveraged previously generated functional kinase activity data for LRRK2 from a robust HEK293 cellular overexpression system, using LRRK2-dependent phosphorylation of Rab10 at threonine 73 as the functional readout and classified variants as activating according to previously established criteria (13,18).

### Building the browser

This browser was developed using R Shiny v1.9.1 and features several widgets, including all *LRRK2* variants identified in the dataset with ANNOVAR annotations and ancestry-specific and full-cohort allele frequencies, visualizations of variant locations in exons and protein domains, kinase activity, and clinical characteristics.

## Data and Code Availability

Data used in the preparation of this article were obtained from the Global Parkinson’s Genetics Program (GP2; https://gp2.org). Specifically, we used Tier 2 data from GP2 release 11 (https://doi.org/10.5281/zenodo.17753486). GP2 data can be requested through AMP PD (https://amp-pd.org). All code generated for this article, and the identifiers for all software programs and packages used, are available on GitHub (https://github.com/GP2code/LRRK2_Browser) and were given a persistent identifier via Zenodo (10.5281/zenodo.20159771).

## Results

Our open-access *LRRK2* Browser contains data on 19,596 variants in *LRRK2* across 11 genetically-determined ancestries. It includes 968 exonic variants, among which 657 are classified as missense and 36 predicted loss-of-function (e.g., frameshift, stopgain, or canonical splicing); 32 variants are kinase-activating, and 14 are currently considered disease-associated (e.g., causal or high-risk) according to MDSGene (https://www.mdsgene.org/), ClinVar (https://www.ncbi.nlm.nih.gov/clinvar/), and/or the Human Gene Mutation Database (HGMD; https://www.hgmd.cf.ac.uk/ac/index.php). The number of unique variants identified differed substantially across ancestries, ranging from 751 variants in the Finnish ancestry group to 9,451 in the European ancestry group, likely reflecting, at least in part, the substantially different sample sizes across ancestries. Excluding the Finnish ancestry group because of its small sample size, the Ashkenazi Jewish ancestry group showed the next lowest number of variants (1,464). Overall, 563 *LRRK2* variants showed global distribution and occurred in every ancestry included here (583 if excluding Finnish ancestry).

For each variant, genetic and functional kinase activity data (where available), together with basic clinical information for identified carriers, were compiled and integrated into an interactive platform. The browser provides comprehensive variant-level annotations, including genomic position, amino acid change, protein domain, predicted functional consequence, CADD score, conservation score, ClinVar clinical significance, and population allele frequencies from gnomAD. A summary of all available information for each variant is provided in **Supplementary Table 1**. In addition, ancestry-specific allele frequencies are displayed for both PD cases and controls within each genetic dataset (**Figure 1A)**.

**Figure 1.**
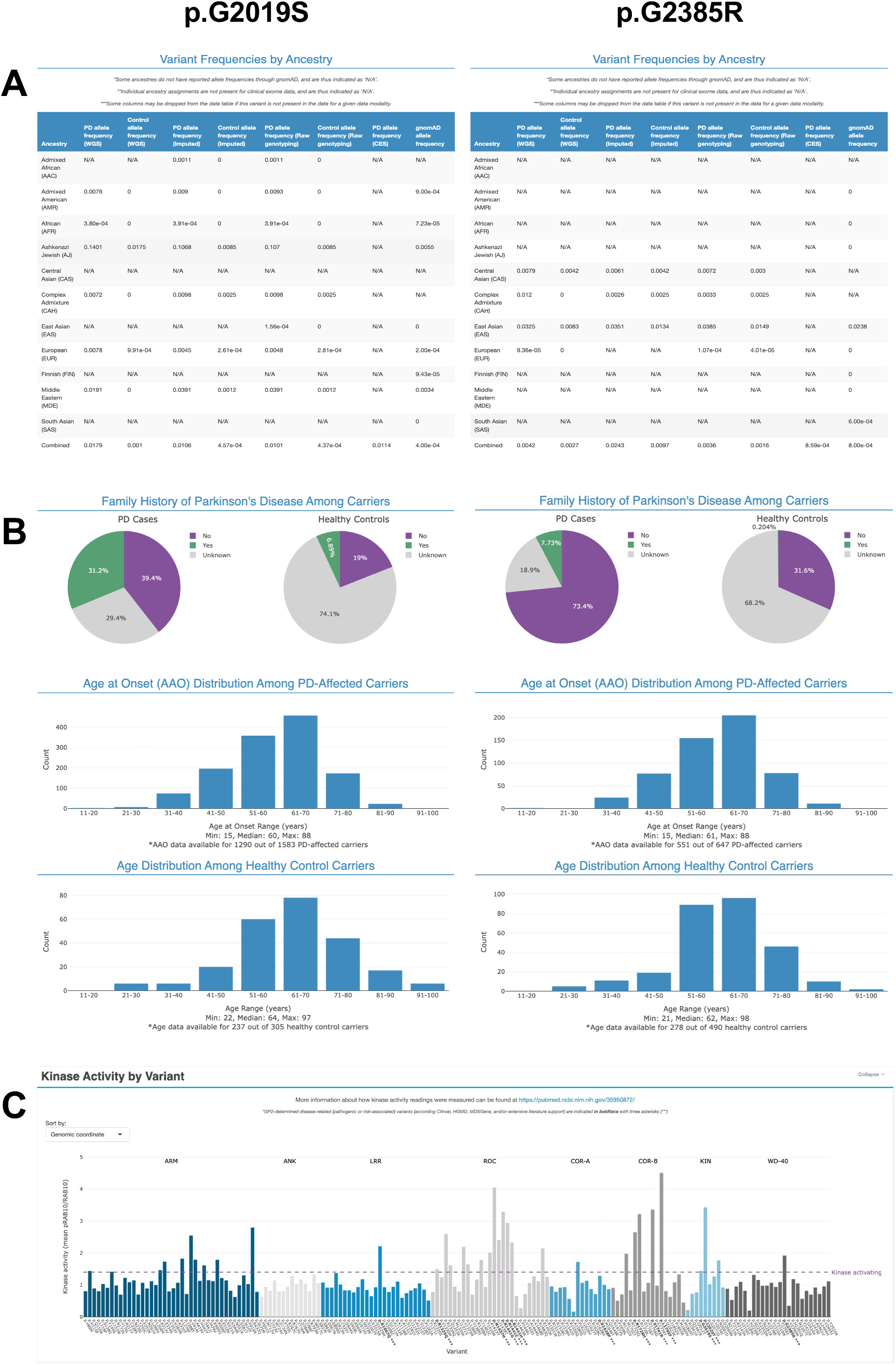
Representative screenshots from the *LRRK2* browser. **(A)** Ancestry-based frequencies of p.G2019S (left) and p.G2385R (right) variants. “N/A” represents lack of data for a given ancestry / genetic dataset pair, or, in the case of gnomAD frequencies, lack of representation for that ancestry in the gnomAD database. **(B)** Clinical information, including family history (top, represented as pie charts for “Yes”, “No”, or “Unknown”) across all carriers of the respective variant, age at onset (middle, represented as a histogram) across PD-affected carriers of p.G2019S (left) and p.G2385R (right) variants, and age (bottom, represented as a histogram) across healthy control carriers. **(C)** Kinase activity (mean pRAB10 / RAB10) for each variant present in the browser with available measurements, plotted as a bar chart.

The browser also provides basic clinical information for variant carriers, including family history of PD, AAO for affected individuals, and age at assessment for unaffected carriers (**Figure 1B**). In addition, kinase activity data are visualized across functional protein domains to facilitate interpretation of variant effects (**Figure 1C**). Summary-level data can be extracted and downloaded for further analysis.

## Discussion

We present the *LRRK2* Browser, an open-access resource focusing on *LRRK2*, linking genetic data with biochemical/functional readouts and providing basic clinical and demographic data on identified variant carriers. The data are derived from GP2’s multi-ancestry datasets, enabling ancestry-specific assessment of *LRRK2* variants. This multi-ancestry representation is particularly important for mitigating European-data-derived bias in variant interpretation and, further, supporting more inclusive clinical trial design, ultimately advancing equitable precision medicine. This is especially relevant as the spectrum of *LRRK2* variants is known to differ substantially across ancestries, with ancestry-enriched variants and founder effects shaping population-specific distributions (26,27). This is prominently exemplified by the marked enrichment of p.G2019S in Ashkenazi Jewish and North African Berber populations, in contrast to its rarity or near absence in Asian and African populations. Integrating these ancestry-specific genetic patterns with functional data is critical for accurate variant interpretation and for informing translational applications.

As *LRRK2* encodes a large, multidomain protein, the browser enables domain-level visualization of variants and directly links these to kinase activity. As shown in **Figure 1C**, many variants located within the ROC, COR-B, and kinase domains are associated with robust increases in LRRK2 kinase activity, consistent with previous reports (13,18). From a clinical perspective, the *LRRK2* Browser provides rapid access to up-to-date, variant-specific information, including pathogenicity classification and relevant clinical data. These include family history of PD in both affected individuals and controls, age at onset in PD carriers, and age distribution among unaffected carriers.

The current *LRRK2* Browser has several limitations. First, while the *LRRK2* Browser leverages the most globally diverse existing PD genetics dataset, European-ancestry samples are nonetheless overrepresented compared to other ancestries. To address this, GP2 continues to generate and release additional genetic datasets with a particular emphasis on currently underrepresented populations. Accordingly, the *LRRK2* Browser infrastructure supports the future integration of forthcoming GP2 datasets, as well as external data from biobanks such as the UK Biobank and All of Us. Notably, despite smaller sample sizes, non-European datasets still represent a valuable resource, comprising over 20,000 PD cases and nearly 20,000 controls across whole-genome sequencing and genotyping platforms. Further, while stringent quality control measures were applied, we acknowledge that genotyping data have not been independently validated, underscoring the importance of using the browser primarily as an exploratory resource for variant interpretation rather than as a definitive dataset for drawing scientific conclusions. Finally, functional interpretation remains incomplete as kinase activity assays are available only for a subset of variants; however, similar to the genetic data, the platform is designed to accommodate future incorporation of additional functional datasets, including expanded kinase activity profiling and other functional readouts such as expression quantitative trait loci (eQTL) analyses.

Altogether, the *LRRK2* Browser enhances the translational potential of *LRRK2* variants for precision medicine by supporting variant risk stratification and informing patient selection strategies. Through its user-friendly interface, the platform is expected to be broadly useful across clinical, biological, biochemical, and genetic disciplines, thereby accelerating the global implementation of LRRK2-targeted therapies.

## Data Availability

All data produced are available in the LRRK2 Browser at http://gp2.org/lrrk2browser

## Acknowledgement

This project was supported by the Global Parkinson’s Genetics Program (GP2; https://gp2.org). GP2 is funded by the Aligning Science Across Parkinson’s (ASAP) (https://ror.org/03zj4c476) initiative and implemented by The Michael J. Fox Foundation for Parkinson’s Research (MJFF) (https://ror.org/03arq3225). For a complete list of GP2 members, see doi.org/10.5281/zenodo.7904831.

This research was supported in part by the Intramural Research Program of the National Institutes of Health (NIH). The contributions of the NIH author(s) were made as part of their official duties as NIH federal employees, are in compliance with agency policy requirements, and are considered Works of the United States Government. However, the findings and conclusions presented in this paper are those of the author(s) and do not necessarily reflect the views of the NIH or the U.S. Department of Health and Human Services.

## Author contributions

LML conceptualized and supervised the study. SMG, VVM, EFT, and MC performed the genetic and clinical data analyses. SMG programmed the browser. ES and DRA performed the functional LRRK2 assays. All authors critically reviewed the browser and contributed to its design and content. SMG, VVM, EFT, MC, and LML drafted the manuscript. All authors reviewed and approved the final version of the manuscript.

